# Socioeconomic and Regional Disparities in Handgrip Strength: A Cross-sectional Analysis of Three Aging Cohorts

**DOI:** 10.64898/2026.09.04.26362242

**Authors:** Rui Li, Sha Nan, Xinyu Feng, Aohan Xu

## Abstract

This cross-sectional study examined the associations of education and body mass index (BMI) with handgrip strength among older adults from the United States, China, and Europe, and quantified education-related inequalities across these populations. We used data from the National Health and Nutrition Examination Survey (NHANES, 2011-2014, n=5,267), the China Health and Retirement Longitudinal Study (CHARLS, 2015, n=16,631), and the Survey of Health, Ageing and Retirement in Europe (SHARE, Wave 6, 2015, n=21,997). Participants were community-dwelling adults aged 50–80 years. Handgrip strength was measured using handheld dynamometers, with maximum values used for analysis. Education was harmonized into three categories (Low, Middle, High). Multivariable linear regression, the Slope Index of Inequality (SII), and association decomposition were employed. European older adults exhibited the highest crude grip strength (mean 34.3 kg), followed by Chinese (31.0 kg) and American (28.8 kg) counterparts. Higher education was significantly associated with greater grip strength in all cohorts. Education-related inequality was greatest in China (SII=4.22, 95% CI: 3.91-4.52) and smallest in the United States (SII=1.67, 95% CI: 1.26-2.08). The educational effect was strongest among European males (3.48 kg) and weakest among Chinese females (0.79 kg). BMI exhibited a suppression pattern rather than mediation, with negative proportional contributions in NHANES (−7.4%) and SHARE (−4.8%). Sensitivity analyses confirmed the robustness of these findings. Considerable cross-national heterogeneity exists in education-related grip strength disparities, with the educational advantage most pronounced in Europe and least effective in China, particularly among women.

## Introduction

Muscle weakness in older age is a critical indicator of physical frailty and sarcopenia, conditions associated with increased risks of disability, chronic illness, cognitive decline, and mortality [1,2]. Handgrip strength, as a simple and reliable measure of overall muscle strength, has become a widely used marker of health status in aging populations [3,4]. Maintaining adequate grip strength is considered essential for functional independence and healthy aging [5]. The prognostic value of grip strength has been demonstrated across diverse populations, with lower levels predicting adverse outcomes including cardiovascular events, falls, and all-cause mortality [3].

The determinants of grip strength in older adults have been extensively investigated. Age and sex are the most consistent predictors, with strength declining progressively after midlife and males exhibiting substantially higher values than females [6–8]. Cross-national normative data have established that grip strength varies considerably across populations, with age-and sex-specific reference values differing by region [6]. Beyond these demographic factors, socioeconomic position (SEP) has emerged as an important correlate. Studies from high-income settings have demonstrated that higher education and income are associated with greater grip strength, independent of age and sex [9,10]. A systematic review confirmed that socioeconomic status, particularly education, is consistently associated with grip strength in older adults [9]. Furthermore, childhood socioeconomic circumstances appear to exert lasting effects on muscle strength in later life, suggesting that the origins of these disparities may extend beyond adult educational attainment [11].

Despite this growing body of evidence, several important gaps remain. First, the majority of existing studies are single-country analyses, which limits our understanding of how social structures, healthcare systems, and cultural contexts shape the relationship between socioeconomic position and muscle health. While some studies have compared grip strength between two populations [11,12], few have simultaneously examined three distinct cohorts representing different economic and social contexts. The United States, China, and Europe differ substantially in population aging trajectories, healthcare provision, and social welfare systems, making comparative analysis particularly informative [13].

Second, the pathways through which education influences muscle strength remain poorly understood. Body mass index has been proposed as a potential mediator, as education may affect nutritional status and body composition [14]. However, the relationship between education, BMI, and grip strength across different populations remains unclear. A systematic review by Harder et al. [14] found that the association between BMI and grip strength varies by age and sex, with lower BMI sometimes associated with lower strength in older populations. It is also unknown whether BMI consistently mediates or potentially suppresses the education-grip strength association in diverse settings.

Third, it remains unclear whether educational gradients in muscle strength differ by gender across national contexts, and in particular, whether women gain equivalent functional advantages from higher education. Recent evidence from China and England suggests that childhood socioeconomic factors may have persistent effects on grip strength trajectories, yet these effects appear to differ by sex and country context [11]. Gender differences in education-related health returns have been observed in other health domains, but less is known about muscle strength specifically. Understanding whether women benefit from education to the same extent as men across different social contexts is essential for designing gender-sensitive public health interventions.

The present study addressed these gaps by examining education-related disparities in grip strength and quantifying education-related inequalities across three large aging cohorts from the United States (NHANES), China (CHARLS), and Europe (SHARE). A secondary aim was to explore the role of BMI in the education-grip strength relationship through association decomposition. By using harmonized measures and consistent analytical approaches across cohorts, we sought to provide comparable estimates of educational disparities in muscle health and to identify whether these disparities vary systematically across populations with different social and policy contexts.

## Materials and Methods

### Data Sources and Study Populations

This cross-sectional study analyzed data from three population-based surveys of community-dwelling older adults.

### NHANES (United States)

The National Health and Nutrition Examination Survey is a nationally representative survey of the non-institutionalized US population. We used data from the 2011-2014 cycles, which included handgrip strength measurements. A total of 5,267 participants aged 50-80 years with complete data on grip strength, age, sex, and education were included.

#### CHARLS (China)

The China Health and Retirement Longitudinal Study is a nationally representative survey of Chinese residents aged 45 years and older. We used the 2015 wave, which conducted physical performance assessments including grip strength. The analytical sample comprised 16,631 participants aged 50-80 years.

#### SHARE (Europe)

The Survey of Health, Ageing and Retirement in Europe is a multidisciplinary and cross-national panel database of Europeans aged 50 and older. We used Wave 6 (2015), harmonized through the easySHARE release. Participants aged 50-80 years from seven European countries (Germany, France, Spain, the Netherlands, Sweden, Denmark) were included (n=21,997).

All surveys obtained ethical approval from their respective institutions, and all participants provided informed consent.

### Handgrip Strength Measurement

Grip strength was measured using handheld dynamometers in all three surveys. In NHANES, a Takei digital dynamometer was used; CHARLS used a Yuejian mechanical dynamometer; SHARE employed a Smedley dynamometer [6]. Each survey measured grip strength multiple times for each hand, and the maximum value (in kilograms) was used for analysis, consistent with standard protocols [6].

Differences in dynamometer brands may introduce minor systematic measurement differences across cohorts, which should be considered when interpreting absolute cross-national differences in grip strength.

### Education Harmonization

Education was classified into three categories based on each survey’s educational attainment measures. For NHANES, Low was defined as less than high school, Middle as high school or some college, and High as college graduate or higher. For CHARLS, Low included illiteracy or primary school; Middle included middle or high school; High included college or above. For SHARE, education was based on ISCED-1997 categories: Low (0-2), Middle (3-4), and High (5-6).

### Body Mass Index

BMI was calculated as weight in kilograms divided by height in meters squared. For NHANES, BMI was derived from physical examination measurements. SHARE provided pre-calculated BMI in the easySHARE dataset. CHARLS did not contain continuous height and weight measures; therefore, decomposition analysis involving BMI was conducted only for NHANES and SHARE.

### Covariates

Age (continuous) and sex (male/female) were included as covariates in all regression models.

## Statistical Analysis

All analyses used unweighted data to ensure cross-cohort comparability, as the three surveys employ non-compatible sampling frames and weight structures.

Linear regression models were fitted to estimate associations of age, sex, cohort, and education with grip strength. Model 1 adjusted for age and sex; Model 2 additionally included cohort; Model 3 further added education.

The Slope Index of Inequality (SII) was calculated within each cohort to quantify education-related disparities in grip strength. The SII quantifies the absolute linear difference in predicted grip strength between individuals at the top versus bottom of the education hierarchy, accounting for the full distribution of educational attainment within each cohort. Higher SII indicates larger absolute educational disparity [15].

Stratified analyses examined education effects (High vs Low) by sex within each cohort.

Association decomposition using the product-of-coefficients method was applied to NHANES and SHARE to quantify the proportional contribution of BMI to the education–handgrip strength association. A negative proportional contribution indicated a suppression pattern.

Primary analyses were implemented separately within each cohort. A pooled regression including a cohort indicator was additionally conducted for descriptive cross-cohort comparison.

## Sensitivity analyses

We conducted three sensitivity analyses to assess the robustness of our findings: (i) excluding participants aged ≥80 years; (ii) excluding the two CHARLS participants aged ≥80 years with high education; and (iii) restricting the CHARLS sample to those with non-missing education. Results remained consistent with the main findings in all three scenarios.

All analyses were performed using R version 4.4.2 (R Foundation for Statistical Computing, Vienna, Austria).

## Results

### Participant Characteristics

Table 1 presents the baseline characteristics of the three cohorts. Participants in CHARLS were slightly younger (mean 61.6 years) compared to NHANES (64.6 years) and SHARE (65.2 years). Grip strength was highest in SHARE (crude mean 34.3 kg), intermediate in CHARLS (31.0 kg), and lowest in NHANES (28.8 kg). Educational distributions differed markedly: SHARE and NHANES had higher proportions of high-education participants (24.6% and 22.6%, respectively) compared to CHARLS (7.4%). Education data were missing for 40.4% of CHARLS participants and 1.3% of SHARE participants, with no missingness in NHANES.

**Table 1.** Baseline Characteristics of Study Participants (Unweighted)

| Characteristic | NHANES<br>(n=5,267) | CHARLS<br>(n=16,631) | SHARE<br>(n=21,997) |
| --- | --- | --- | --- |
| Age, mean (SD) | 64.6 (9.5) | 61.6 (7.9) | 65.2 (7.8) |
| Male, % | 48.4 | 49.8 | 46.4 |
| Grip strength, kg, mean<br>(SD) | 28.8 (9.0) | 31.0 (9.8) | 34.3 (11.7) |
| Education, % |  |  |  |
| Low | 13.0 | 28.6 | 41.7 |
| Middle | 64.4 | 23.6 | 32.4 |
| High | 22.6 | 7.4 | 24.6 |
| Missing | 0.0 | 40.4 | 1.3 |

**Footnotes:** Values are unweighted means (SD) or percentages. SHARE sample limited to Wave 6 participants aged 50–80 years from seven European countries.

### Age-Related Decline in Grip Strength

Figure 1 illustrates the age-related decline in handgrip strength across the three cohorts. In all three populations, grip strength decreased progressively with age. European older adults maintained higher grip strength throughout the 50–80 age range, while American older adults exhibited the lowest levels. The divergence between cohorts widened with advancing age: at age 50, the difference between SHARE and NHANES was approximately 1.0 kg; by age 80, this gap had widened to approximately 4.5 kg. The Chinese cohort occupied an intermediate position throughout the age range.

**Figure 1.**
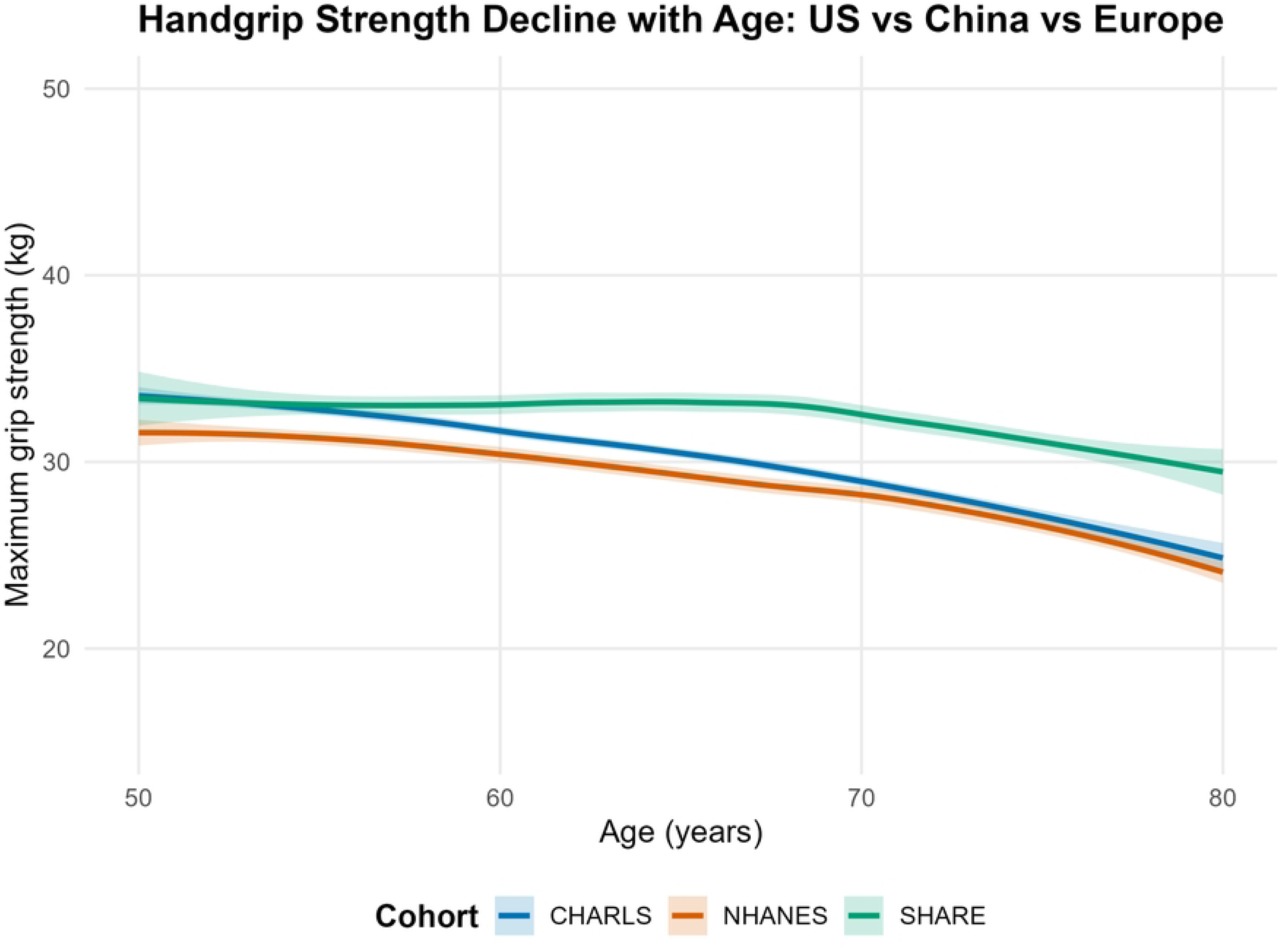
Handgrip strength trajectories by age in the US (NHANES), China (CHARLS), and Europe (SHARE). Solid lines represent loess-smoothed means; shaded bands indicate 95% confidence intervals. SHARE was subsampled (20%) for visualization due to its large sample size. The y-axis shows maximum grip strength in kilograms.

### Multivariable Regression

Table 2 presents the linear regression results examining the independent associations of age, sex, cohort, and education with grip strength. In the fully adjusted model (Model 3), each additional year of age was associated with a 0.368 kg lower grip strength (p < 0.001). Female sex was associated with 15.184 kg lower strength (p < 0.001). Compared to CHARLS, NHANES participants had 1.796 kg lower grip strength, and SHARE participants had 4.788 kg higher grip strength (both p < 0.001). Education showed a graded association: Low education was associated with 2.522 kg lower grip strength than High education, and Middle education with 0.666 kg lower strength (both p < 0.001). The model explained 57.9% of the variance in grip strength.

**Table 2.**
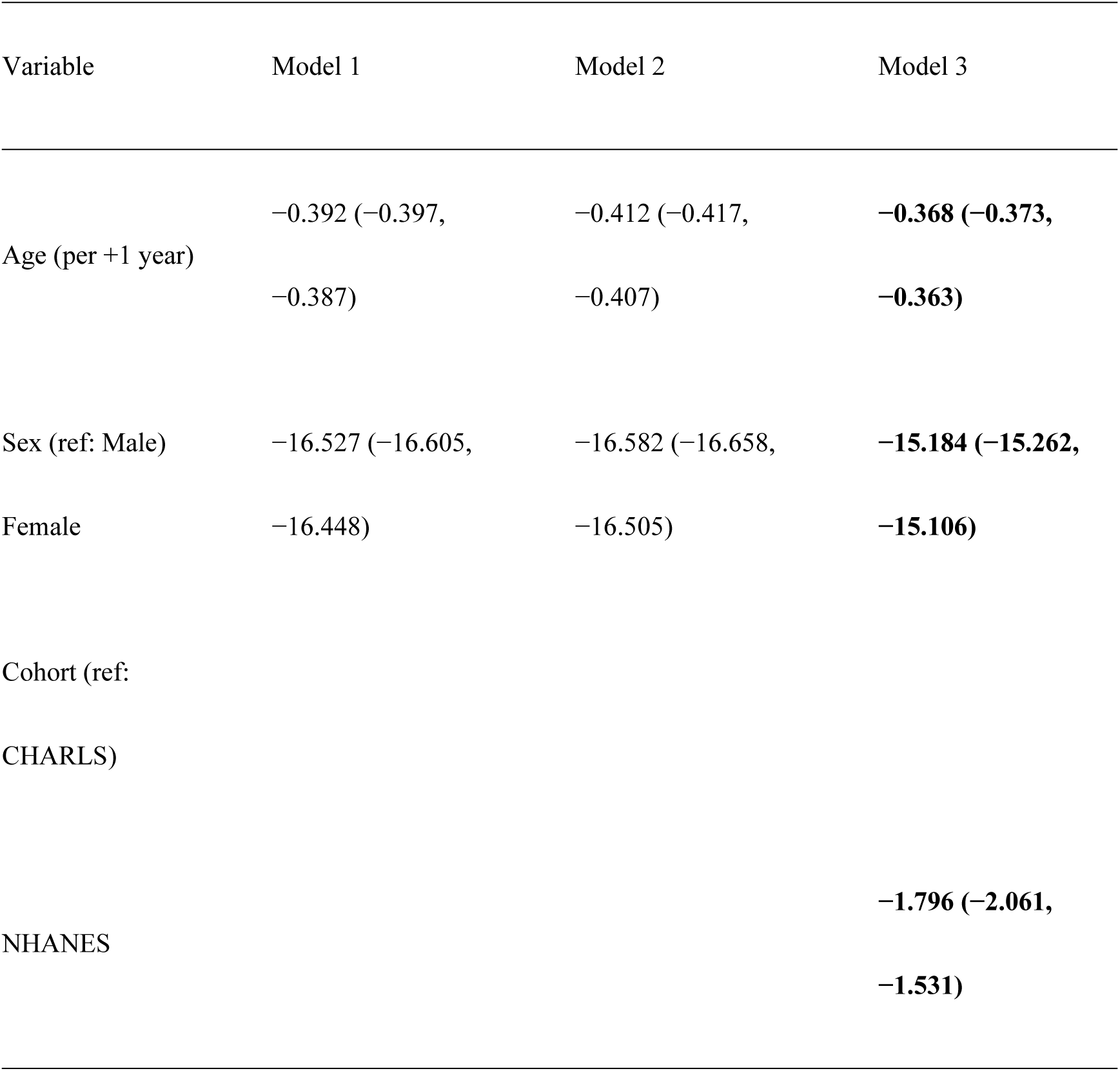
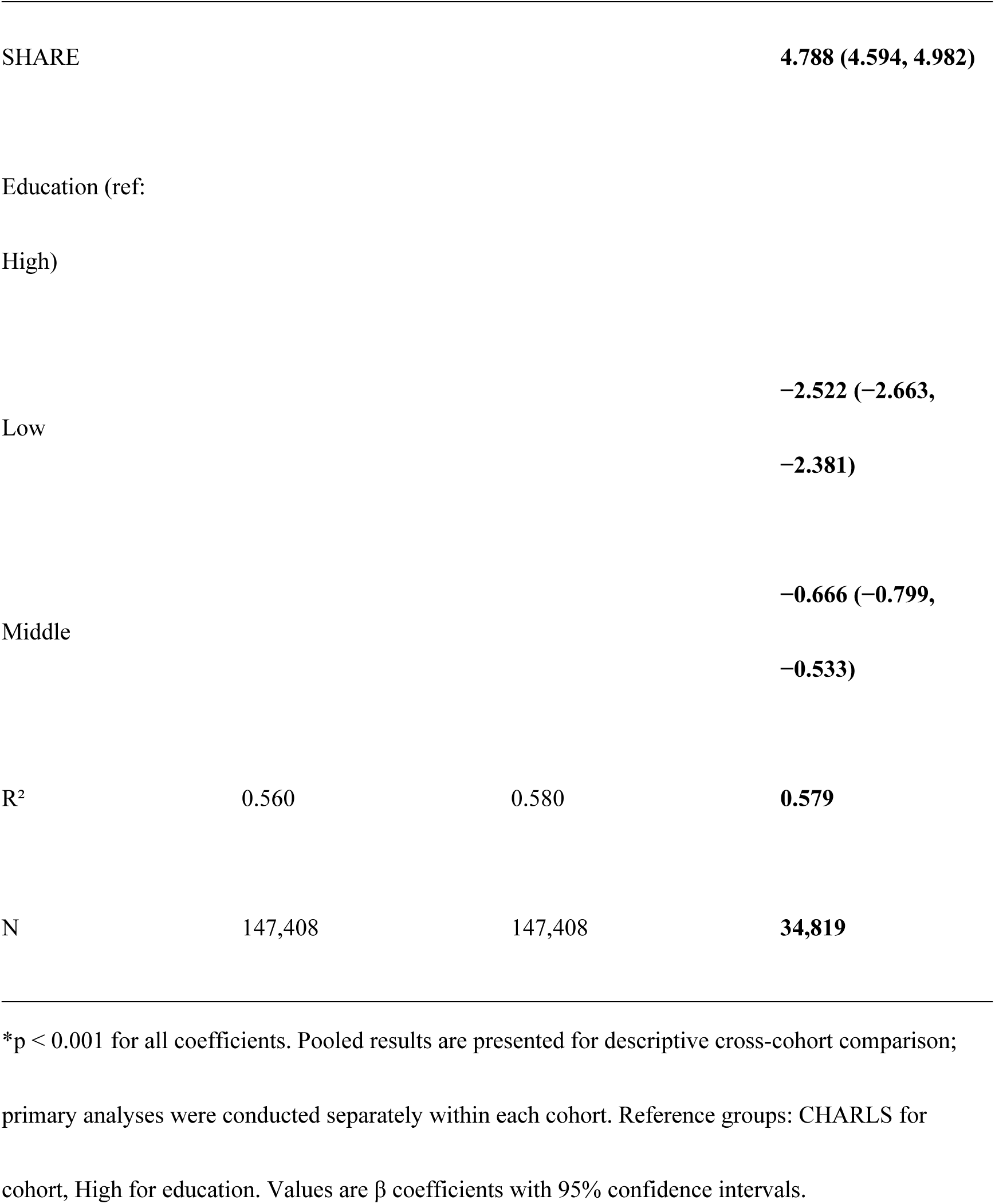
Linear Regression Models for Grip Strength (kg) – Pooled Descriptive Analysis.

Figure 2 displays the Slope Index of Inequality for education-related grip strength disparities across the three cohorts. Education-related inequality was greatest in CHARLS (SII=4.22, 95% CI: 3.91-4.52), intermediate in SHARE (SII=2.77, 95% CI: 2.57-2.96), and smallest in NHANES (SII=1.67, 95% CI: 1.26-2.08). The confidence intervals did not overlap between CHARLS and the other two cohorts, indicating that the education-grip strength gradient was significantly steeper in China than in the United States or Europe.

**Figure 2.**
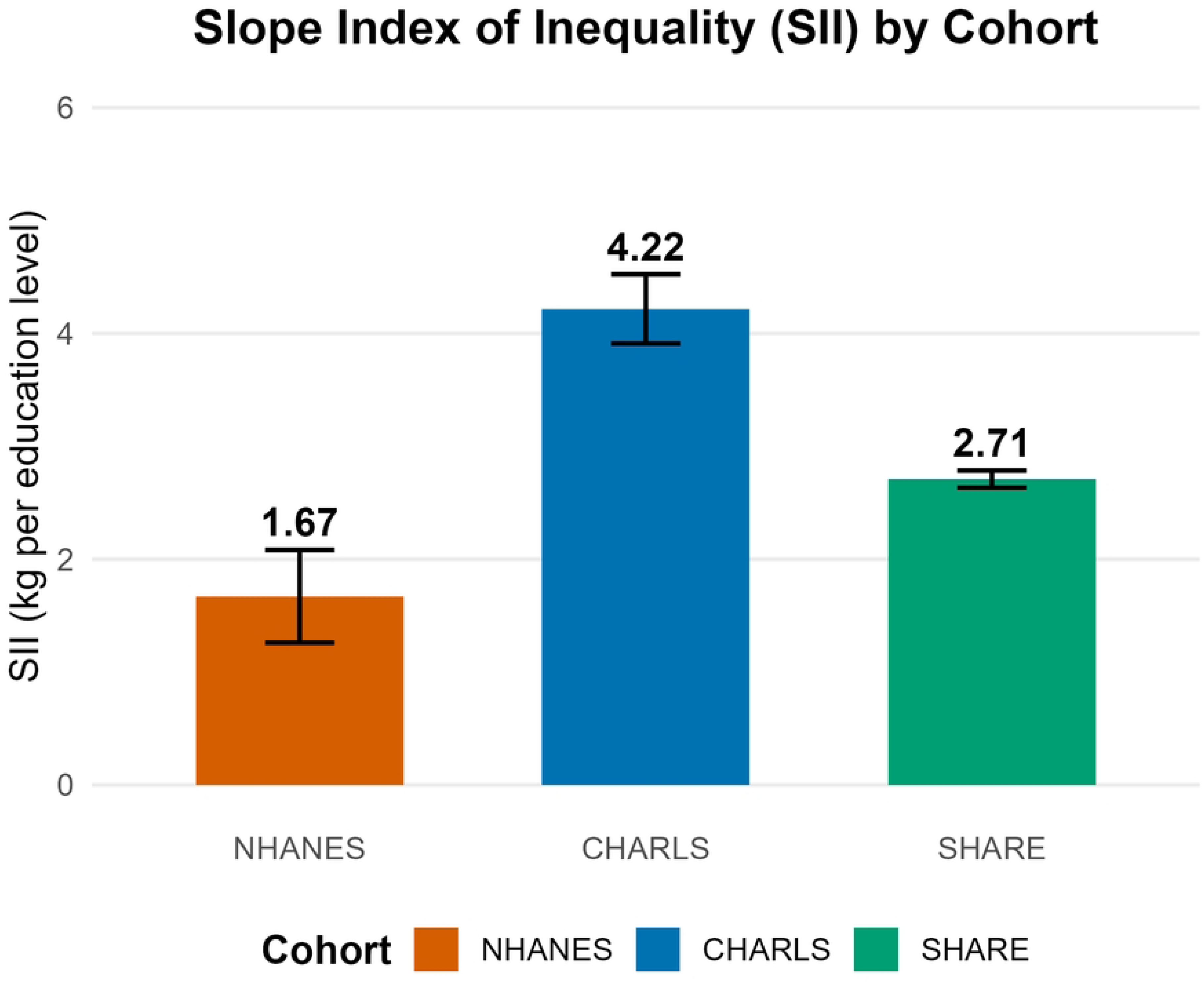
Slope Index of Inequality (SII) for education-related grip strength disparities across cohorts. Bars represent SII values (kg per education level); error bars indicate 95% confidence intervals. Higher SII values indicate greater education-related inequality in grip strength.

### Stratified Analyses by Sex and Cohort

Table 3 displays education effects (High vs Low) stratified by sex within each cohort. The education effect was consistently larger in males than females across all three cohorts. The largest sex difference was observed in CHARLS, where the education effect was 1.84 kg for males but only 0.79 kg for females. The education effect was strongest among SHARE males (3.48 kg) and weakest among CHARLS females (0.79 kg). The sex difference in educational effect was smallest in SHARE (0.79 kg) and largest in CHARLS (1.05 kg). The forest plot summarizing these stratified effects is provided in Supplementary Figure 2.

**Table 3.**
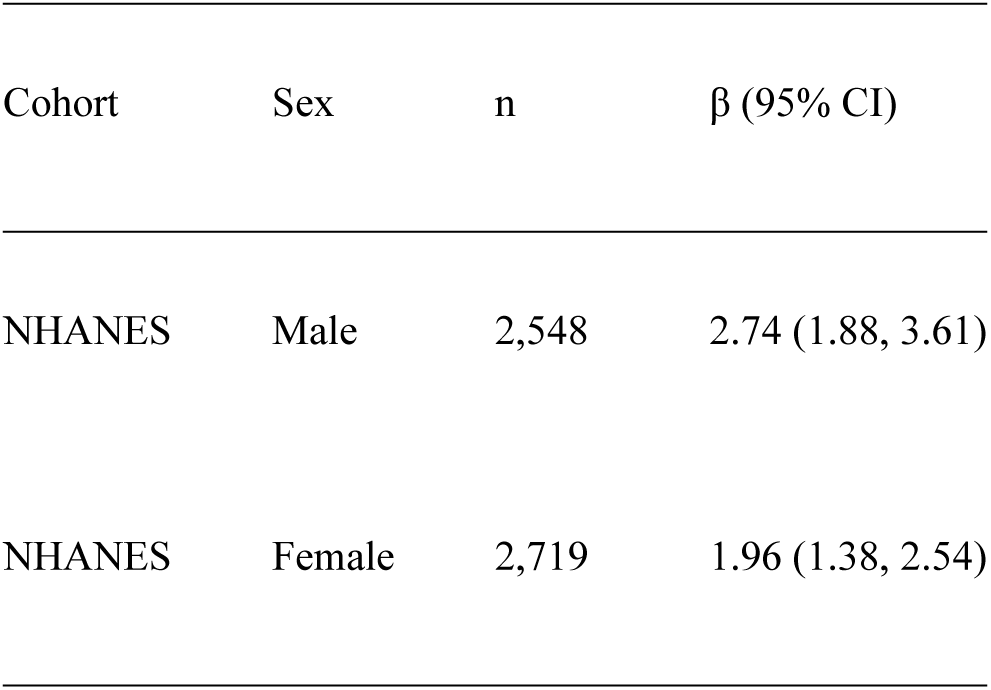
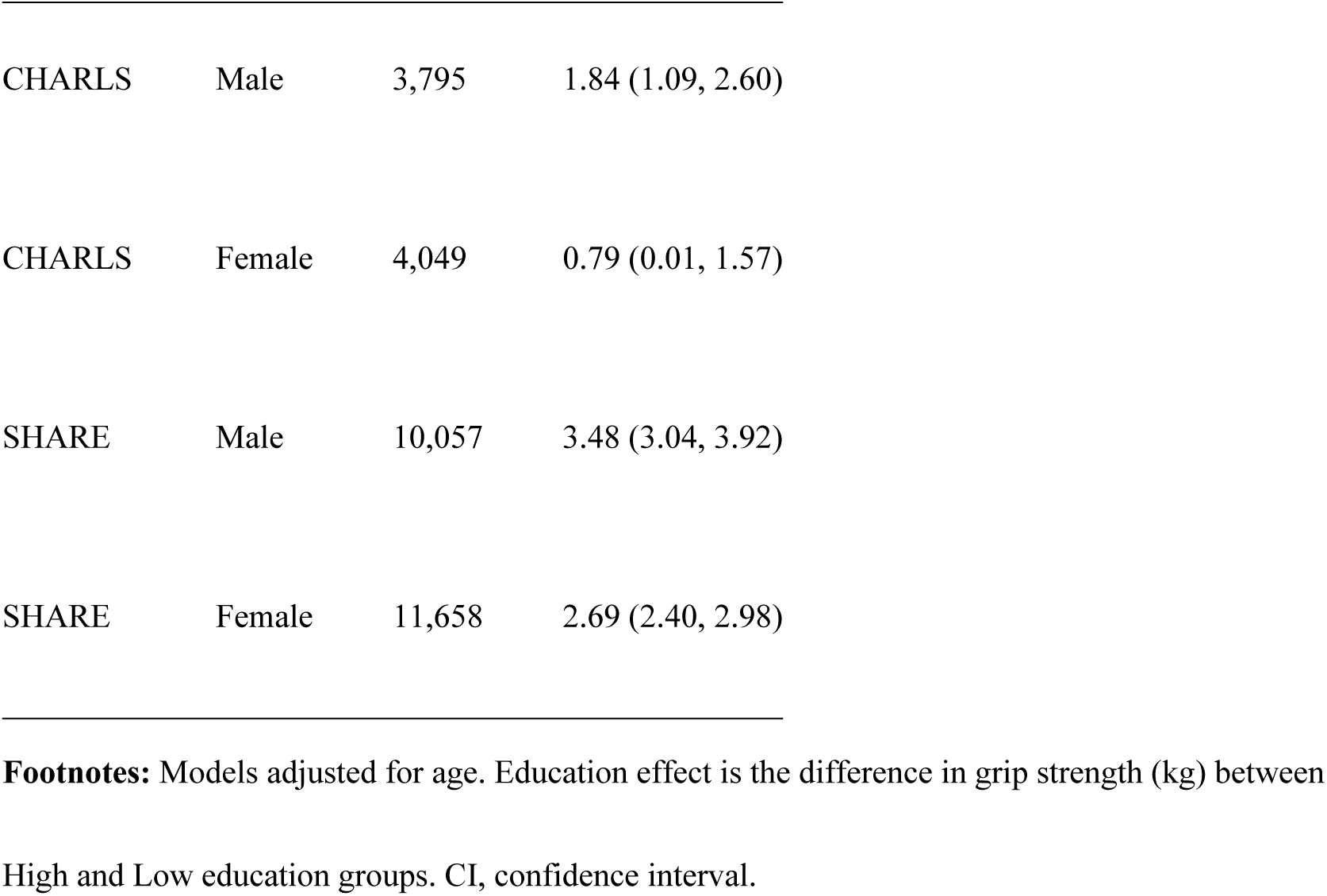
Education Effects (High vs Low) by Sex and Cohort (Cohort-Specific Models)

### Interaction Effects

The interaction model examining whether the education effect varied significantly across cohorts is presented in Supplementary Table 1. The CHARLS × High education interaction term was negative (β=-1.784, p < 0.001), indicating that the education effect in China was significantly smaller than in the United States. Conversely, the SHARE × High education interaction term was positive (β=0.835, p=0.020), indicating that the education effect in Europe was significantly larger than in the United States. These findings confirm that the educational gradient in grip strength is steepest in Europe, intermediate in the United States, and flattest in China.

### Association Decomposition: Role of BMI

The association decomposition examining the proportional contribution of BMI to the education-grip strength association is presented in Supplementary Figure 1 and Supplementary Table 2. In NHANES, the proportional contribution of BMI was negative (−7.4%), indicating a suppression pattern. In SHARE, similar patterns were observed, with a proportional contribution of −4.8%. These findings suggest that higher education was associated with lower BMI in both cohorts, and lower BMI was associated with lower grip strength, resulting in a net negative proportional contribution. This suppression pattern indicates that BMI counteracts part of the education-related strength advantage rather than acting as an intermediate pathway.

## Sensitivity Analyses

Sensitivity analyses confirmed the robustness of the main findings (Supplementary Table 3). When excluding participants aged ≥80 years, all core coefficients remained stable (age:-0.365 vs-0.368 in the main model; education High: 2.541 vs 2.522). Excluding the two CHARLS participants aged ≥80 years with high education produced nearly identical estimates (education High: 2.523 vs 2.522). Restricting the CHARLS sample to those with non-missing education also yielded consistent results. The direction, magnitude, and statistical significance of all core associations remained unchanged across all three sensitivity scenarios.

## Summary of Results

In summary, this study found that European older adults had the highest grip strength, followed by Chinese and American counterparts. Education was significantly associated with grip strength in all cohorts, but the magnitude of this association varied considerably, with the strongest effects observed in Europe and the weakest in China. Education-related inequality in grip strength, as measured by SII, was greatest in China and smallest in the United States (Figure 2). Sex-stratified analyses revealed that the educational effect was consistently larger in males, with Chinese females showing the smallest educational advantage (Table 3). BMI exhibited a suppression pattern rather than a mediating role in the education-grip strength association (Supplementary Table 2). These findings were robust across multiple sensitivity analyses (Supplementary Table 3).

## Discussion

This cross-national study of older adults from the United States, China, and Europe yielded three principal findings. First, European older adults exhibited the highest grip strength, American older adults the lowest, with the Chinese cohort positioned intermediately. Second, education was significantly associated with grip strength in all cohorts, but the magnitude of this association varied substantially, with the strongest effects observed in Europe and the weakest in China. Third, education-related inequality in grip strength, quantified by SII, was greatest in China and smallest in the United States, with the sex-stratified analyses revealing that Chinese females derived the least functional advantage from higher education.

### Cross-National Differences in Grip Strength

The higher grip strength observed in the European cohort is consistent with previous international comparisons [6,12]. European populations generally exhibit higher levels of physical activity in daily life, more favorable nutritional profiles, and more comprehensive healthcare systems compared to the United States [20]. The relatively lower grip strength in the American cohort aligns with concerns about obesity prevalence and sedentary lifestyles in the US population [17]. The intermediate position of the Chinese cohort may reflect a balance between higher daily physical activity levels and lower socioeconomic resources among older adults [18]. Notably, the divergence between cohorts widened with age (Figure 1), suggesting that the cumulative effects of these environmental and lifestyle factors become more pronounced in later life.

### Education-Related Inequality in Muscle Health

The finding that education-related inequality in grip strength was greatest in China (SII=4.22) warrants particular attention. This pattern is consistent with broader evidence that educational inequalities in health are more pronounced in settings with historically limited educational expansion and persistent regional disparities [19]. Older Chinese adults came of age during periods when educational opportunities were heavily constrained by geographic location, family background, and political upheaval [19]. These historical circumstances created large and persistent gaps in educational attainment that continue to shape health outcomes in later life.

The intermediate position of Europe in our SII analysis (2.77) aligns with evidence that European welfare states partially mitigate health inequalities through universal healthcare, social protection, and active aging policies [20,26]. The smaller educational gradient in the United States (1.67), despite higher income inequality, may reflect the universal availability of certain health resources or the influence of other factors such as obesity that reduce the health advantage of higher education [21]. These findings suggest that the relationship between education and muscle strength is not fixed but is shaped by the broader social and policy context.

### Gender Disparities and Life Course Origins

The weak educational effect among Chinese females (0.79 kg) is particularly notable. This finding is consistent with evidence that educational returns in health-related domains are smaller for women in China, reflecting historical gender disparities in educational access, labor force participation, and caregiving responsibilities [10]. In contrast, the educational effect among European females (2.69 kg) was substantially larger, suggesting that more equitable social structures and gender policies in Europe enable women to convert educational attainment into health benefits more effectively [22].

Life course evidence further contextualizes these findings. A recent study using CHARLS and the English Longitudinal Study of Ageing found that childhood socioeconomic position—including parental education and childhood health—was independently associated with grip strength trajectories in midlife and older age [11]. In the CHARLS cohort, participants whose parents were illiterate had grip strength that was, on average, 0.36 kg lower. This suggests that the educational disparities we observed in older Chinese adults may have roots in early life circumstances, with adult educational attainment representing both individual achievement and the accumulation of childhood disadvantage.

### The Suppression Pattern of BMI

The negative proportional contribution of BMI to the education-grip strength association (−7.4% in NHANES, −4.8% in SHARE) indicates a suppression pattern rather than mediation. Higher education was associated with lower BMI in both cohorts, and lower BMI was associated with lower grip strength, producing a net negative contribution. This is consistent with observations that underweight and low body mass may be detrimental to muscle strength in older populations [23]. Rather than mediating the education effect, BMI appears to exert a countervailing influence that partially masks the true educational advantage. When BMI is accounted for, the total educational gradient becomes amplified, suggesting that weight management strategies should be considered in the context of preserving muscle strength, rather than focusing on weight alone.

### Clinical Implications and Future Directions

For clinicians and public health practitioners, these findings suggest several actionable directions. First, asking about educational background may help identify older adults at higher risk for muscle weakness, particularly among Chinese women. Second, context-specific interventions in China should focus on improving health literacy and promoting physical activity among less-educated older adults, especially women. Third, weight management programs for older adults should incorporate resistance exercise and adequate protein intake to preserve muscle mass while addressing excess weight [24]. Fourth, the persistence of educational gradients even in universal healthcare settings highlights the need to address health literacy and health behaviors beyond access to care alone.

### Limitations

This study has several limitations. The cross-sectional design precludes causal inference. Education classification was harmonized but may not capture population-specific differences in educational quality. A large proportion (40.4%) of CHARLS participants lacked educational data; the primary analysis relied on complete-case data, and although sensitivity analyses suggested no strong association between missingness and grip strength, selection bias cannot be fully excluded. BMI data were unavailable in CHARLS, limiting the decomposition analysis to NHANES and SHARE. Differences in dynamometer brands may introduce minor systematic measurement differences across cohorts [6]. Finally, this study focused on crude and age-sex-adjusted educational disparities to enhance cross-country comparability; uniformly harmonized lifestyle and chronic disease variables were unavailable across all three surveys, and residual confounding cannot be excluded.

### Strengths

This study has several strengths, including the use of three large, population-based cohorts with harmonized variables, the application of SII to quantify educational inequality, the association decomposition analysis to examine the role of BMI, and the robustness of findings confirmed by multiple sensitivity analyses.

## Conclusions

In conclusion, this study provides robust evidence that education-related inequalities in grip strength are substantial and variable across cohorts. The largest disparities were observed in China, and the weakest educational effects among Chinese females suggest that gender-specific interventions may be needed. BMI does not mediate but rather suppresses the education-grip strength association in the cohorts where it could be examined. Future longitudinal studies with harmonized measures of education, BMI, and physical activity are needed to further elucidate the mechanisms underlying these cross-national differences.

## Declarations

### Authors’ contributions

Rui Li and Sha Na contributed equally to this work. RL: Conceptualization, Data curation, Formal analysis, Writing – original draft. SN: Data curation, Validation, Writing – review & editing. XF: Validation, Writing – review & editing. AX: Validation, Writing – review & editing. All authors read and approved the final manuscript.

## Data Availability

All data used in this study are publicly available from third-party repositories. NHANES data are available at https://www.cdc.gov/nchs/nhanes. CHARLS data are accessible at https://charls.charlsdata.com. SHARE data are available at https://share-eric.eu. All three datasets require registration and approval for access. The authors did not generate any new data for this study. The R code used for all analyses is provided in the Supplementary Material.

https://charls.charlsdata.com

https://www.cdc.gov/nchs/nhanes

## Acknowledgements

The authors thank the investigators and participants of CHARLS, NHANES, and SHARE for making these data publicly available. We also thank the data collection teams and funding agencies that supported these studies.

## Data availability statement

All data used in this study are publicly available. CHARLS data are accessible at https://charls.charlsdata.com. NHANES data are available at https://www.cdc.gov/nchs/nhanes. SHARE data are available at https://share-eric.eu. The R code used for all analyses is provided in the Supplementary Material.

## Ethics approval statement

This study utilized publicly available, de-identified data from three population-based cohorts. All original studies received ethical approval from their respective institutional review boards, and all participants provided informed consent. As this was a secondary analysis of existing anonymized data, no additional ethical approval was required.

## Patient consent statement

Not applicable. This manuscript contains no individual person’s data in any form.

## Supplementary Materials

The following supplementary materials are available online:

• **Supplementary Table 1.** Interaction Effects: Cohort × Education (Pooled Model)

• **Supplementary Table 2.** Association Decomposition: Proportional Contribution of BMI

• **Supplementary Table 3.** Sensitivity Analysis Summary

• **Supplementary Figure 1.** Path Diagrams for BMI Decomposition

• **Supplementary Figure 2.** Forest Plot of Education Effects by Cohort and Gender

## References

1. Cruz-Jentoft AJ, Bahat G, Bauer J, et al. Sarcopenia: revised European consensus on definition and diagnosis. Age Ageing. 2019;48(1):16–31.

2. Dodds RM, Syddall HE, Cooper R, et al. Grip strength across the life course: normative data from twelve British studies. PLoS One. 2014;9(12):e113637.

3. Leong DP, Teo KK, Rangarajan S, et al. Prognostic value of grip strength: findings from the Prospective Urban Rural Epidemiology (PURE) study. Lancet. 2015;386(9990):266–273.

4. Bohannon RW. Hand-grip dynamometry predicts future outcomes in aging adults. J Geriatr Phys Ther. 2008;31(1):3–10.

5. Rantanen T, Era P, Heikkinen E. Maximal isometric grip strength and mobility among 75-year-old men and women. Aging Clin Exp Res. 1994;6(2):133–139.

6. Dodds RM, Syddall HE, Cooper R, et al. Global variation in grip strength: a systematic review and meta-analysis of normative data. Age Ageing. 2016;45(2):209–216.

7. Stevens PJ, Syddall HE, Patel HP, Martin HJ, Cooper C, Sayer AA. Is grip strength a good marker of physical performance among community-dwelling older people? J Nutr Health Aging. 2012;16(9):769–774.

8. Alley DE, Shardell MD, Peters KW, et al. Grip strength cutpoints for the identification of clinically relevant weakness. J Gerontol A Biol Sci Med Sci. 2014;69(5):559–566.

9. Beller J, Geyer S. The relationship of socioeconomic status and grip strength: a systematic review. J Epidemiol Community Health. 2017;71(10):1028–1034.

10. Lei X, Shen Y, Smith JP, Zhou G. Sibling gender composition’s effect on education: evidence from China. J Popul Econ. 2017;30(2):569–590.

11. Chisala M, Hardy R, Cooper R, Li L. Associations of childhood socioeconomic position and health with trajectories of grip strength from middle to older ages in populations from China and England. Maturitas. 2025;191:108154.

12. Tyrovolas S, Koyanagi A, Olaya B, et al. Factors associated with skeletal muscle mass, sarcopenia, and sarcopenic obesity in older adults: a multi-continent study. J Am Med Dir Assoc. 2016;17(11):1071.e13-1071.e19.

13. Gu D, Dupre ME, Warner DF, Zeng Y. Changing health status and health expectancies among older adults in China: gender differences from 1992 to 2002. Soc Sci Med. 2009;68(12):2170–2179.

14. Harder K, Hossein M, Lang J, et al. Body mass index and grip strength in older adults: a systematic review. BMC Geriatr. 2023;23:833.

15. Moreno-Betancur M, Latouche A, Menvielle G, et al. Relative index of inequality and slope index of inequality: a review of the methods. Int J Epidemiol. 2015;44(4):1399–1418.

16. Zeng Y, Gu D, Purser J, Hoenig H, Christakis N. Associations of environmental factors with elderly health and mortality in China. Am J Epidemiol. 2010;172(12):1399–1410.

17. Gu D, Hsin PL. Obesity and physical function in older adults: evidence from the Health and Retirement Study. J Aging Health. 2016;28(6):1069–1092.

18. Zeng Y, Gu D, Land KC. A new method for correcting the underestimation of disabled life expectancy in the oldest old in China. Demography. 2007;44(3):533–551.

19. Zhou X, Wang G. Educational inequality in China: a review of the literature. Chin J Sociol. 2017;3(2):234–256.

20. Avendano M, Berkman LF, Brugiavini A, Pasini G. The long-run effect of social security on health: evidence from pension reform in Europe. J Health Econ. 2014;37:89–103.

21. Singh GK, Siahpush M, Hiatt RA, Timsina LR. Dramatic increases in obesity and overweight prevalence and body mass index among ethnic-immigrant and social class groups in the United States, 1976-2008. J Community Health. 2011;36(1):94–110.

22. Avendano M, Glymour MM, Banks J, Mackenbach JP. Health disadvantage in US adults aged 50 to 74 years: a comparison of the health of rich and poor Americans with that of Europeans. Am J Public Health. 2009;99(3):540–548.

23. Prado CM, Wells JC, Smith SR, Stephan BC, Siervo M. Sarcopenic obesity: a critical appraisal of the current evidence. Clin Nutr. 2012;31(5):583–601.

24. Kim YJ, Moon S, Yu JM, Chung HS. Implication of diet and exercise on the management of age-related sarcopenic obesity in Asians. Geriatr Gerontol Int. 2022;22(9):695–704.

25. Pongiglione B, Beller J, Kelleher J, et al. Cross national patterns in educational inequalities in functional limitations among middle aged and older adults at two time points. SSM Popul Health. 2024;28:101725.

26. Solé-Auró A, Lozano M, Renteria E, et al. Health across Generations in Europe. The role of Age, Gender, and Education. J Popul Ageing. 2026. doi:10.1007/s12062-026-09456-2.

